# Absorption and Co-expression Modules Show Where Polygenic and Proteomic Risk Scores Diverge in Neurodegenerative Diseases

**DOI:** 10.64898/2026.08.24.26361271

**Authors:** Charles Zheng, Manu Shivakumar, Li Shen, Dokyoon Kim

**Author notes:** This author is the corresponding author. These authors contributed equally to this work (Charles Zheng, Manu Shivakumar).

## Abstract

Polygenic and proteomic risk scores are both proposed for pre-symptomatic stratification, yet the extent to which they provide overlapping or complementary information has not been measured across neurodegenerative disease. To quantify this overlap, we define absorption as the fraction of a polygenic score’s predictive contribution accounted for by an out-of-fold proteomic score and estimate it among 9,434 to 9,820 UK Biobank participants. Absorption did not track *h*^2^, as Alzheimer’s with *APOE*, Alzheimer’s without *APOE* and Parkinson’s carried matched SNP *h*^2^ of 0.068, 0.061 and 0.069 yet absorbed 0.73, 0.39 and 0.19, with amyotrophic lateral sclerosis at 0.22. Residual genetic signal remained in all four, indicating that proteomic risk scores did not fully capture the predictive information contained in polygenic risk scores. The proteins associated with a polygenic score and the proteins a proteomic score selects overlap no more often than chance, converging only where *APOE* dominates. Across 33 plasma co-expression modules built in 41,358 disease-free participants, germline signal concentrates in modules rather than spreading, with the summary component of seven modules associated with the score for Alzheimer’s with *APOE* and none for Parkinson’s, even though Parkinson’s carries the lysosomal genetic architecture that the lysosomal module M32 encodes. This module has 89% of its members associated with AD germline risk yet none was used by the proteomic score. Rebuilding the co-expression modules in *All of Us* gave an adjusted Rand index of 0.582 against the 0.686 attainable within that cohort, and 29 of 33 modules stayed together above a permutation null. Cross-cohort transferability was predictable from module coherence in the discovery cohort, supporting the reuse of this module partition as a fixed reference dictionary.

## 1. Introduction

Two stratification methods have been proposed and implemented for estimating patient risk before disease onset. The first uses the germline, static genome, summarized as a polygenic risk score (PRS). The second uses proteomic readings in plasma and sometimes cerebrospinal fluid (CSF), which represents the patient’s state of health and physiology more dynamically and is now assayable at scale by affinity platforms.^1,2^ With multi-modality data becoming increasingly available, clarifying how much one modality substitutes for the other becomes essential for determining whether PRS and ProRS provide redundant or complementary predictive information, and whether both are needed for risk stratification.

Recent work by Woerner *et al*. showed that proteomic risk scores (ProRS) trained on prevalent samples often surpassed PRS in predictive performance on incident cases while retaining complementary information, but also observed that when used in conjunction, the fraction of PRS’s contribution after conditioning on ProRS scaled with heritability of the predicted trait.^3^ However, Woerner *et al*. did not specifically analyze, on a protein or pathway resolution, how the two scores overlap or substitute for each other, nor did they individually categorize neurodegenerative diseases such as Alzheimer’s Disease (AD), Parkinson’s disease (PD), amyotrophic lateral sclerosis (ALS), and frontotemporal dementia (FTD).

Previous studies have shown that *APOE ε*4 carriage leaves a reproducible imprint on the circulating proteome.^4–6^ This presents evidence that plasma proteome readings, which builds the ProRS, can be influenced by genomic variants, and individual proteins can be predicted by effects of single loci and thus, upon aggregation, PRS. Therefore, we ask the question for each of AD, PD, ALS and FTD: to what extent can ProRS can account for proportion of genetic risk explained by the PRS signal, and can this be explained by the individual proteins that they each act on to predict disease risk?

Proteomic and polygenic stratification may converge on the same biological processes without involving the same individual proteins. For example, genetically associated and disease-predictive proteins may occupy different roles within one pathway, and many individually weak associations may become detectable only when treated as a coordinated system. Co-expression modules provide such a higher-order representation by capturing biologically coherent processes rather than isolated proteins. Extensive literature has shown that protein modules recover reproducible biology and remain coherent across cohorts and tissues.^7–10^ Prior serum work further demonstrated that modules associated with incident disease and mortality were subject to both cis- and trans-genetic regulation.^11^ We therefore ask whether the predictive redundancy observed between PRS and ProRS reflects sparse associations across the plasma proteome or concentration within a limited number of co-expression modules, and whether the two scores converge at the module-level despite limited overlap among individual proteins.

We address these questions in UK Biobank, using 2,920 Olink analytes, polygenic scores built exclusively from non-UK-Biobank discovery GWAS, and 41,358 healthy plus incident-case participants for the proteome-wide work. Our module-level analysis is validated in 9,211 disease-free participants of *All of Us*, assayed on a later Olink panel. Our contributions are fourfold. We quantify the extent to which proteomic risk captures the predictive contribution of polygenic risk as an effect size with a bootstrap interval, and find it substantial for AD, partial in every disease examined. We then show that the proteins ProRS selects and the ones polygenic risk can affect are largely different populations, overlapping no more often than chance except for AD with APOE. We then analyzed 33 data-derived co-expression modules and observed the two sets’ modular separation as well as patterns of their respective enrichment. Finally, we ask whether those module definitions are stable across cohort transfer by rebuilding them in All of Us and measuring their agreement with UK Biobank, which yields a reproducible module membership dictionary for later studies relating germline or proteomic risk to the plasma proteome.

## 2. Methods

### 2.1. Cohorts, phenotyping and quality control

Discovery analyses used UK Biobank,^12,13^ restricted to participants of European genetic ancestry with Olink Explore 3072 proteomics at the baseline instance.^1^ Cases were defined from linked hospital and death records by ICD-10 code (AD G30, PD G20, ALS G12.2, FTD G31.0) and classified as prevalent or incident relative to venepuncture. Three distinct analysis populations are used. The nested logistic models use the complete-case intersection of proteome, genotype and covariates with a downsampled control pool, giving 9,434–9,820 participants per disease. The association analyses use every participant with genotype data but without a recorded diagnosis at venepuncture, including the incident diagnoses, giving 38,602–39,807 per disease. Modules are built once in the 41,358 participants free of all four diagnoses with proteomic data. Covariates throughout were age, age squared, sex and the first ten genotype principal components.

Modules and absorption were assessed in an external cohort. *All of Us* contributed 9,969 participants with Olink Explore HT plasma proteomics.^14,15^ Of the 2,920 UK Biobank module proteins, 2,819 were present on Explore HT by gene symbol and survived QC. Proteins missing in more than 20% of participants and participants missing more than 20% of proteins were dropped, and the remainder were standardized and mean-imputed, whereas the UK Biobank matrix was *k*-nearest-neighbour imputed.

### 2.2. Polygenic risk scores

To preclude sample-overlap inflation, discovery summary statistics were taken exclusively from non-UK-Biobank GWAS (Table 1).^16–19^ Posterior effect sizes were obtained with PRS-CS under the continuous-shrinkage prior against the 1000 Genomes European LD reference,^20^ and scores computed with PLINK.^21^ Two AD scores were constructed, one genome-wide and one excluding chr19:44.4–46.5 Mb (GRCh37),^22^ with the exclusion applied to the summary statistics before shrinkage so that the resulting posterior is *APOE* -free by construction. SNP heritability on the liability scale was estimated by LD score regression.^23^

**Table 1.** Study design. Phenotype counts are for European-ancestry UK Biobank participants with both Olink plasma proteomics and genome-wide genotypes (N = 38,670 complete cases). SNP-heritability is on the liability scale from LD score regression.

| Trait | ICD-10 | Discovery GWAS | Prevalent | Incident | Controls | Liability $h^2$ (SE) |
| --- | --- | --- | --- | --- | --- | --- |
| AD, with *APOE* | G30 | Kunkle 2019 | 101 | 483 | 42,545 | 0.068 (0.011) |
| AD, no *APOE* | G30 | Kunkle 2019 | 101 | 483 | 42,545 | 0.061 (0.009) |
| PD | G20 | FinnGen R13 | 105 | 652 | 42,372 | 0.069 (0.012) |
| ALS | G12.2 | van Rheenen 2021 | 36 | 220 | 42,873 | 0.028 (0.003) |
| FTD* | G31.0 | Ferrari 2014 | 20 | 69 | 43,040 | 0.026 (0.022) |

\*FTD is unstable (the standard error of $h^2 \approx$ the estimate) and is excluded from subsequent claims.

### 2.3. Proteomic risk scores

ProRS^3^ were fitted by LASSO logistic regression with `glmnet`,^24^ with proteins penalized and covariates forced in unpenalized, and *λ* selected by inner cross-validation. A repeated stratified five-fold design was used in which all prevalent cases enter every training fold while incident cases and a downsized control sample are scored strictly out of fold; the entire procedure was repeated three times with distinct seeds, giving fifteen selection opportunities per protein. Because the ProRS is never exposed to the genetic score, one cached ProRS per disease is compared against every genetic score. A protein counts as ProRS-selected when it is selected in at least 7 of the 15 folds. As a sensitivity analysis, every ProRS was refitted by elastic net at *α* = 0.5 with the penalty being the only difference.

### 2.4. Proteomic absorption of polygenic predictive signal

For each disease we fitted four nested logistic models on an identical complete-case set, these being covariates alone, covariates plus PRS, covariates plus ProRS, and both.^3^ Using Δ*R*^2^ increment,^25^ absorption is

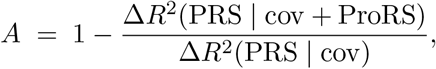

the proportion of the polygenic score’s own predictive contribution accounted by the proteome. Confidence intervals came from a case-control-stratified bootstrap with 2000 resamples, and between-disease comparisons use a paired bootstrap that resamples the shared control pool once and applies the same resample to both diseases.^26^ The DeLong test on the area under the curve is also reported.^27^

### 2.5. PRS-associated proteins

A protein was called PRS-associated if its abundance tracked that score among participants without a recorded diagnosis at proteomic assessment, including controls and incident cases. Prevalent cases were excluded so that established disease, its treatment, or its physiological consequences could not alter protein abundance and be misattributed to a PRS association. Each of the 2,920 assayed proteins was regressed individually on the standardized PRS with age, age^2^, sex and PC1–10 as covariates, and the resulting *p*-values were Benjamini–Hochberg corrected at *q <* 0.05 within each score, and every subsequent *q* value carries the same correction. Two genomic regions are reported separately from the rest. The major histocompatibility complex and the 17q21.31 inversion^1,28,29^ were associated with a very wide range of traits and contain strong *cis*-pQTLs, so every polygenic score carried weight there and every score therefore appeared to associate with the proteins those regions encode. Proteins encoded in the two regions are held out of the proteome-share denominator for that reason and counted separately (Fig. 1a).

**Fig. 1.**
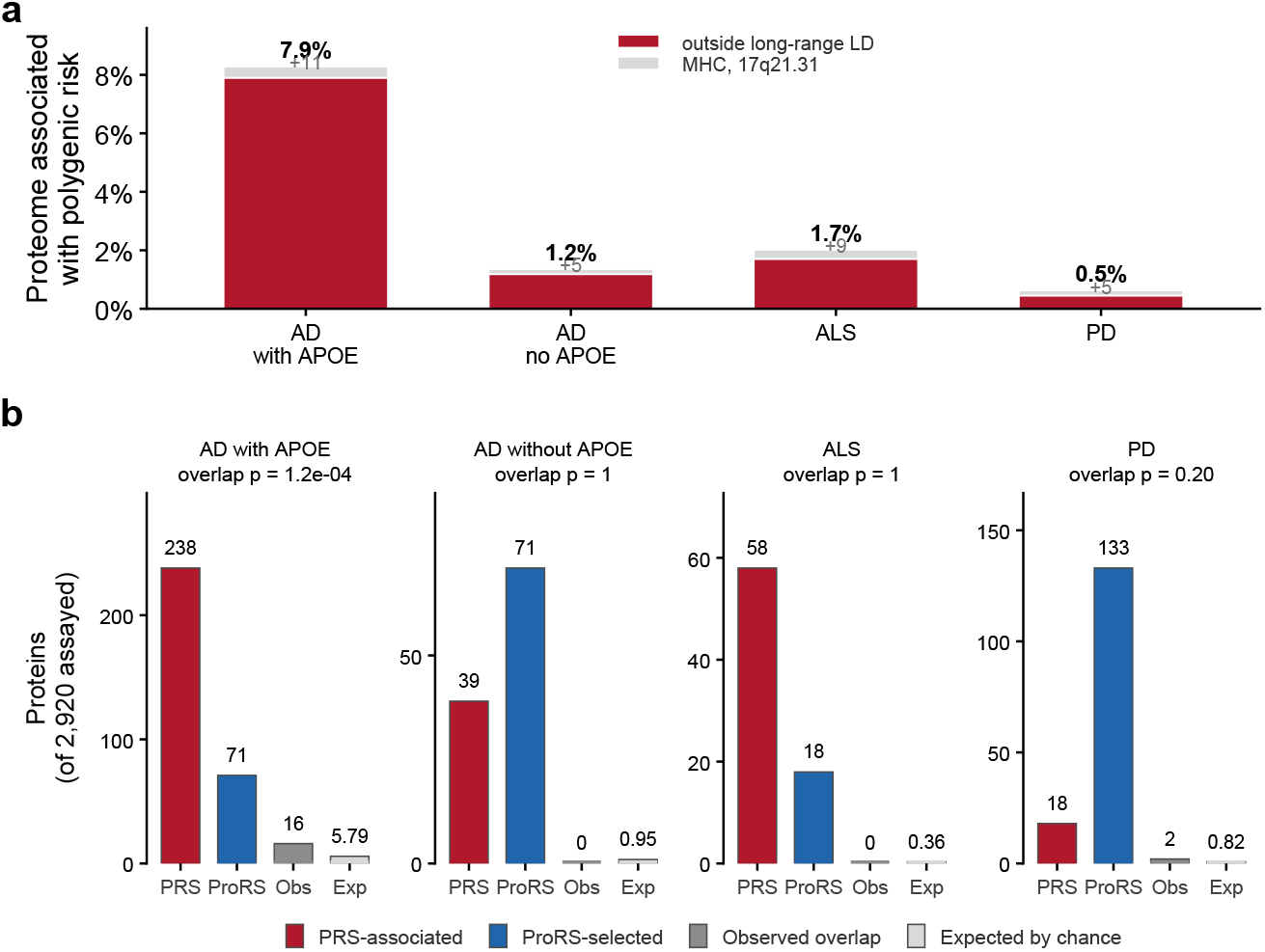
Two protein populations, in 38,602 to 39,807 participants without a recorded diagnosis at venepuncture, depending on trait. (a) Share of the assayed proteome each polygenic score is associated with at q < 0.05, over the 2,873 analytes outside the MHC and the 17q21.31 inversion, with the count contributed by those two regions drawn as a separate segment. (b) Absolute count of PRS-associated proteins, ProRS-selected proteins at 7 of 15 folds, their observed overlap and the overlap expected by chance, with the one-sided hypergeometric p for excess overlap above each panel.

### 2.6. Plasma proteome co-expression modules

In the 41,358 disease-free participants, each protein was residualized on the covariate design by ordinary least squares, retaining residuals only. Residuals were standardized and Pearson-correlated pairwise, and the signed distance 1 *− r* clustered by Ward’s minimum-variance criterion following established practice.^7^ The hierarchical clustering merged at each step the pair of clusters producing the smallest increase in within-cluster sum of squares, recording the merges in a tree-like dendrogram, which was used to cut at twelve candidate resolutions spanning *K* = 8 to 50 clusters. Each resolution was characterized on reproducibility, measured by partitioning participants at random into halves and scoring each half’s partition against the full-data partition by the adjusted Rand index (ARI) over five independent splits, as well as the smallest module the cut produced (Fig. 2c). Reproducibility rises monotonically across the sweep and so cannot select a resolution on its own; thus, we selected for *K* = 35 as the finest resolution before the smallests module size degrades from a plateu of 17 to 8 (Supplementary Data). Each module was summarized by its eigenprotein, the score on the first principal component of its members’ residualized abundances, which assigned one value to every participant. Two modules whose eigenproteins correlate closely are carrying the same signal under two names, so any pair correlating above 0.85 was merged. The retained cut requested 35 clusters, and two such merges returned the 33 modules analyzed throughout (Fig. 2c).

**Fig. 2.**
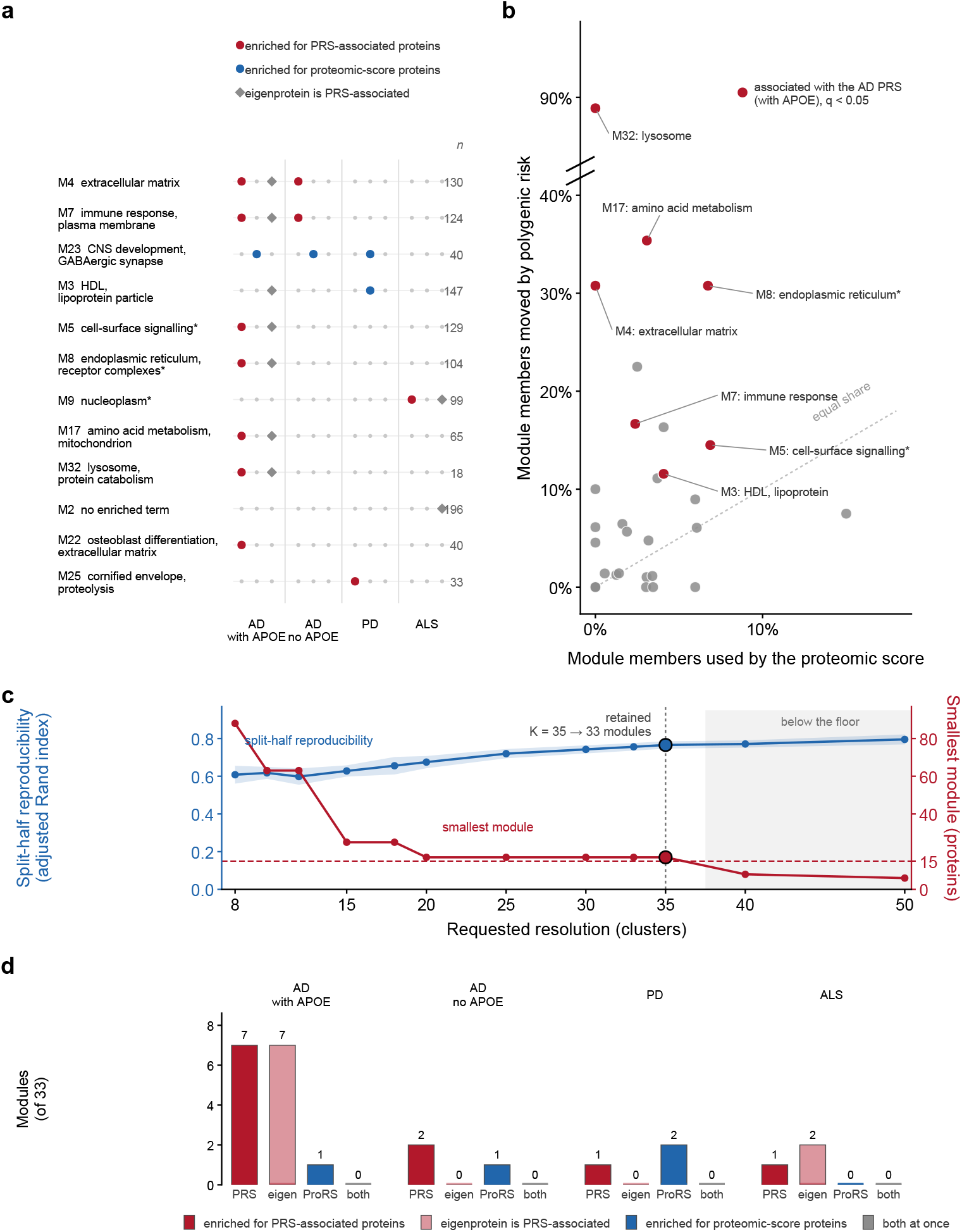
Module-resolution analysis. Modules are defined once in the 41,358 participants free of all four diagnoses; the association axes are evaluated in each disease’s own analysis population. (a) The 12 modules significant on at least one of the three axes, of the 33 tested. An asterisk marks a module named by its most enriched gene-ontology term where that term does not clear q < 0.05. (b) Per module, fraction of ProRS-selected proteins against the fraction of AD with APOE PRS-associated proteins, all 33 modules, filled red where the eigenprotein itself can be predicted by AD PRS. (c) Split-half reproducibility and smallest module size across twelve resolutions. (d) Barplot showing the counts of how many modules have eigenprotein associated with PRS, is enriched for ProRS-selected proteins (in t ≥ 7/15 folds), is enriched for PRS-associated proteins, and is enriched for both ProRS and PRS proteins.

Modules were annotated by hypergeometric over-representation of Gene Ontology terms with the assayed panel as background. Module-level association was assessed by regressing each module’s standardized eigenprotein on the standardized polygenic score with the same covariates, in disease-free participants only so that disease-consequence cannot confound any discovery. Each module was additionally tested for over-representation of the PRS-associated set and of the ProRS-selected set by hypergeometric test against the assayed panel, giving three per-module axes in total.

### 2.7. Cross-cohort comparison

Module transfer was assessed in *All of Us* using proteins shared with UK Biobank. The two cohorts were residualized and partitioned independently through the same clustering procedure, and agreement was quantified by the adjusted Rand index (ARI). As an internal reference ceiling, *All of Us* was split in half ten times, each pair of half-sample partitions was compared, and the median ARI was retained. Module-specific preservation was assessed by the fraction of each UK Biobank module’s protein pairs assigned to the same *All of Us* module. This co-location fraction was compared with 2,000 size-matched random protein sets, converted to a *z*-score, and tested using one-sided *p*-values with Benjamini–Hochberg correction across 33 modules. In a complementary analysis, UK Biobank module membership was held fixed without reclustering *All of Us*. Module coherence, defined as the variance explained by the first principal component, was recomputed in *All of Us* and compared with the corresponding UK Biobank values.^30^ UK Biobank modules were derived in European-similar disease-free participants, whereas the *All of Us* analysis included all ancestries because restricting to the 2,681 European-similar participants would reduce the available sample from 9,211 by approximately fourfold.

Absorption was additionally evaluated in *All of Us* by applying the frozen UK Biobank ProRS weights without refitting. Refitting was not attempted because only 23 to 43 disease cases were available, so the transferred score tests the UK Biobank weights rather than the *All of Us* proteome’s best achievable fit.

## 3. Results

### 3.1. Proteomic substitution for polygenic risk is partial in every disease and its magnitude tracks one locus

Adding ProRS to the covariates model improved discrimination in every disease and by a wider margin than what PRS achieved, with AD rising from an area under the curve of 0.819 for covariates alone to 0.855 with PRS and 0.903 with ProRS (Table 2), echoing *Woerner et al*.’s findings. Absorption was 0.727 (95% CI 0.645–0.806) for the *APOE* -containing AD score, 0.394 (0.119–0.667) for AD with *APOE* excluded, 0.221 (0.009–0.509) for ALS and 0.187 (0.001–0.380) for PD. The residual polygenic increment’s interval excluded zero in all four contrasts, with Δ*R*^2^(PRS | ProRS) at 0.0226 (0.0134–0.0341), 0.0057 (0.0014–0.0128), 0.0081 (0.0016–0.0184) and 0.0047 (0.0013–0.0101) for AD with APOE, without APOE, ALS and PD respectively.

**Table 2.**
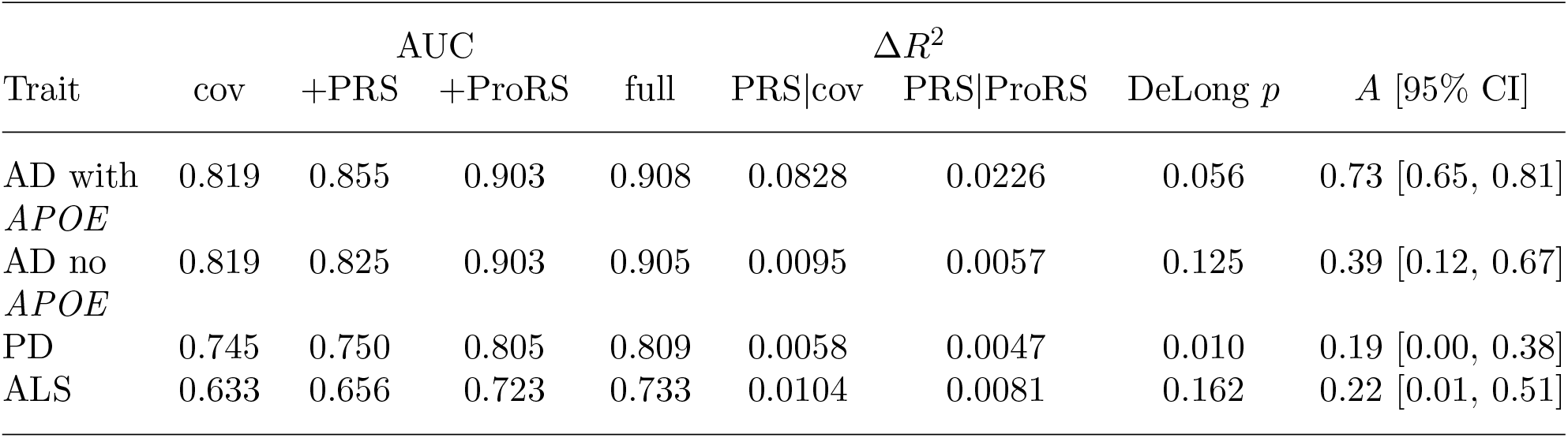
The four-model AUC and absorption results. ΔR^2^ values are Nagelkerke increments. DeLong p reflects AUC significance of PRS given ProRS and covariates. Absorption intervals are percentile bootstrap (B = 2000, 3 cross-validation repeats). FTD is characterised in Table 1 but omitted here, its absorption denominator being below the stability threshold.

AD with APOE had the greatest absorption of any disease model at 0.73 whereas PD was the lowest at 0.19, and SNP heritability alone cannot explain the differences between them, since liability-scale *h*^2^ is 0.068, 0.061 and 0.069 for the AD, AD-without-*APOE* and PD scores, all within one standard error of each other (Table 1). Different disease absorption values were calculated based on different disease populations and PRS increments; therefore, the within-disease contrast between AD with and without APOE are more technically sound where the outcome, analysis population, covariates, and ProRS were held constant. Excluding the APOE region reduced absorption from 0.73 to 0.39, isolating the APOE region as a major contributor to the unusually high absorption observed for AD.

An attempt to repeat the analysis in *All of Us* reproduces the point estimates but cannot address the contrast. Absorption for the *APOE* -containing AD score was 0.470 (0.248–0.862) on 39 prevalent cases, an interval overlapping the UK Biobank estimate, and PD gave 0.249 in the European-similar subset, likewise overlapping (Supplementary Data). AD without *APOE* was not estimable there, its denominator Δ*R*^2^(PRS | cov) falling to about 5 *×* 10^*−*5^. The AD-versus-PD contrast rests on 20 and 35 cases in that subset and is under-powered, so this replicates the point estimates and explicitly not the dissociation. The frozen UK Biobank proteomic scores transferred to *All of Us* without refitting, giving an AUC of 0.865 for AD and 0.791 for PD (Supplementary Data).

### 3.2. The proteins polygenic risk associates with and the proteins the proteomic score uses are generally distinct

Regressing every assayed protein on each PRS in disease-free participants demonstrates the plasma’s association with genomic signal on an individual-protein resolution. The *APOE* - containing AD score was associated with 7.9% of the proteome (227 of 2,873 proteins), against 1.2% for AD’s polygenic background, 1.7% for ALS and 0.5% for PD (Fig. 1a). Proteins encoded in the MHC and in the 17q21.31 inversion are held out of that denominator and drawn separately, so percentages are computed over the 2,873 analytes outside those regions while the counts described below are over all 2,920. AD’s polygenic background associates with the products of its own risk genes^16^ (*TREM2 q* = 2.5 *×* 10^*−*9^, *CD33 q* = 3.1 *×* 10^*−*5^, *CR1 q* = 3.7 *×* 10^*−*3^), whereas the PD score associates with neither of its main risk genes *SNCA* (*q* = 0.87) nor *GBA* (*q* = 0.85);^31^ *LRRK2* is not represented on the Olink panel and was not tested.

AD with APOE is the outlier here as it was for absorption. Its PRS associates with the most plasma proteins at 238, and it is the only model whose overlap with the ProRS-selected set exceeds chance, at 16 shared proteins against 5.79 expected (Fig. 1b). The three other models are largely disjoint. AD without *APOE* and ALS have separate ProRS, PRS sets, and PD shares two against 0.82 expected, which is not statistically distinguishable from chance. PD has the smallest PRS-associated footprint at 18 proteins, and its low absorption further suggests that inherited PD risk leaves only a limited detectable signature in the plasma proteome.

### 3.3. Germline signal and the proteomic score’s vocabulary separate at module resolution

Unsupervised clustering of the residualized proteome in UK Biobank yielded 33 co-expression modules after two pairs being merged from 35 modules, ranging from 17 to 717 proteins. Modules recovered recognizable biology, including immune cell-surface signalling (*q* = 4 *×* 10^*−*23^), lysosomal hydrolases (*q* = 3 *×* 10^*−*23^), sarcomere (*q* = 6 *×* 10^*−*10^) and lipoprotein particles (*q* = 2 *×* 10^*−*3^) (Fig. 2a). Each module was analyzed for whether its membership was enriched for PRS-associated proteins, whether it was enriched for ProRS-selected proteins (selected in *t ≥* 7*/*15 folds), and whether its eigenprotein was associated with PRS (Fig. 2a). No module was enriched for both the ProRS-selected and the PRS-associated set in any disease, indicating that the two sets stay separate at the network resolution as well as at protein resolution (Fig. 2d).

For PD, while 2 modules were enriched for ProRS-selected proteins, 1 was enriched for PRS-associated proteins, and no module’s eigenprotein was PRS-associated. This is expected, however, as PD only has 18 PRS-associated proteins to begin with. This contrasts with AD with APOE, where 7 modules were enriched for PRS-associated proteins and 7 module eigen-proteins were themselves associated, dominated by the 18-protein lysosomal module M32, 89% of whose members are individually PRS-associated. The two sets of seven overlap in six modules, M4, M5, M7, M8, M17 and M32, which are enriched for PRS-associated members and also associated as a module eigenprotein with PRS, but this pattern dissociates once *APOE* is removed from the score.

Although LASSO may shrink correlated proteins and thus potentially reduce the number of ProRS, PRS protein overlap, their separation here does not depend on the penalty. Refitting every ProRS by elastic net enlarged the selected sets by roughly half, yet the overlap grew no faster than chance, the observed-to-expected ratio for AD with *APOE* moving from 2.76 to 2.22 while the other three models stayed at or below chance, and no module was enriched for both sets under either penalty (Supplementary Data).

### 3.4. Module definitions transfer across cohort, and coherence tracks which ones

Upon defining *K* = 33 modules in UK Biobank, two ways to define module construction in *All of Us* at *K* = 33 were tested. First, modules were built *de novo* in *All of Us* on 2,819 shared proteins in 9,211 disease-free participants gave an adjusted Rand index of 0.582 against the UK Biobank partition. Read against the internal ceiling of 0.686, the median over ten random split-half rebuilds within *All of Us* (range 0.628 to 0.779), that recovers 84.8% of the attainable agreement (Fig. 3a).

**Fig. 3.**
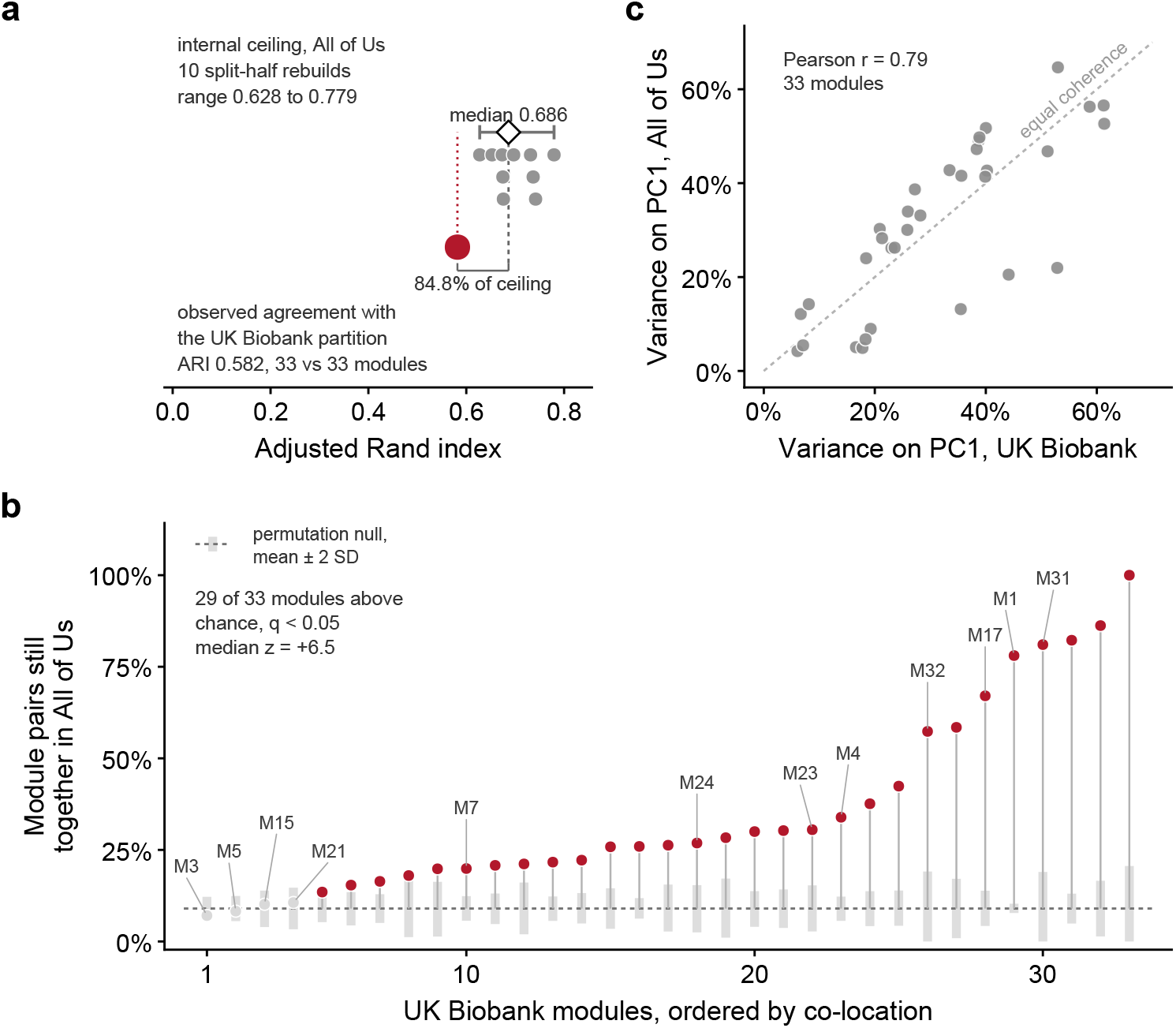
Transfer of the UK Biobank module definition to All of Us. All of Us partitioned **independently** from UK Biobank at K = 33: (a) Partition agreement between the two cohorts with the ten split-half rebuilds drawn individually and module counts annotated; (b) Per module, the fraction of within-module protein pairs still assigned together by the de novo All of Us partition, against a size-matched permutation null from the same 2,819 shared proteins. All of Us partitioned **identically** as UK Biobank: (c) Variance on the first principal component, membership held at the UK Biobank definition, measured once in each cohort.

Under the independent partitions, co-location was also tested to determine whether the members of one UK Biobank module are still partitioned together in *All of Us*, and 29 of 33 modules beat a size-matched permutation null at *q <* 0.05, with a median *z* of +6.5 (Fig. 3b). The lysosomal module M32 keeps 57.4% of its member pairs together against a chance level near 9%, a *z* of +9.57. M3, the HDL and lipoprotein module, is one of the four failures at *z* = *−*1.19.

The second approach applied UK Biobank’s *K* = 33 partition unchanged to *All of Us* and asked whether the modules are equally coherent there. Comparing the variance on the first principal component of the same 33 modules across the two cohorts gave a Pearson correlation of 0.79. Coherence is therefore stable across these two Olink-assayed cohorts. Since both use the same assay platform, this does not establish that modules are free of technical structure, but it does show that internal consistency is reproducible rather than being particular to one sample. (Fig. 3c).

## 4. Discussion

Proteomic substitution for polygenic risk is never complete, and its size is not explained by heritability alone. Three scores whose liability-scale heritability agrees within one standard error give absorption estimates spanning most of the available range, ordered by whether the score carries a large-effect locus whose product is itself measured in plasma.^32^ APOE is heavily implicated in our findings: the absorption gap, the proteome-wide footprint and the single lysosomal module all belong to *APOE*.

M32 is a readout of lysosomal biology, holding 18 proteins across four enzyme classes, cathepsins, glycosidases, sphingolipid enzymes and peptidases, together with *ARSA* and *CREG1*. A plasma lysosomal signature of patients carrying *APOE ε*4 has been reported,^6^ and lysosomal *ABCA1* placed downstream of cholesterol accumulation in *APOE4* models supplies possible a mechanism.^33^ Our addition is the 18-protein module membership that was arrived at having no reference to any gene set while still being concentrated for lysosomal functions relevant to APOE and AD genetic risk. On the other hand, PD inverts the expectation that pathway relevance predicts plasma legibility. Robak *et al*. found a genome-wide excess of damaging lysosomal storage disorder gene variants in Parkinson’s cases, confirmed *GBA* and *SMPD1*, and reported 56% of cases carrying at least one such variant.^34^ *SMPD1* is a member of M32. And yet, the disease with the lysosomal architecture associates with neither M32 nor *GBA* nor any of the 33 eigenproteins, and the disease dominated by a lipid transport locus associates with M32 the most. Although previous work predicted PD up to 14 years before diagnosis from 2,937 plasma proteins, we did not find a strongly detectable germline-driven plasma signature for PD.^35^

M23 was consistently concentrated for ProRS-selected proteins across AD and PD. Its gene-ontology annotation places it in central nervous system development and the GABAergic synapse, and several members are already established fluid markers of neurodegeneration, including NEFL for neuroaxonal damage,^36^ GFAP for astroglial injury,^37^ and SYT1 and NPTXR among synaptic markers.^38^ The remainder spans myelin (MOG, OMG, KLK6), perineuronal matrix (BCAN, NCAN), and synaptic adhesion and vesicle cargo (SEZ6, SEZ6L, SLITRK1, VGF, CHGB, SCG2, SCG3). AD and PD were associated with this module through different proteins. AD selected BCAN, GFAP, NPTXR, SYT1 and VGF, PD selected F3, LRTM2, NCAN, PTPRN2, SCG2 and SLITRK1, and NEFL was the only member both scores use. This agreement between AD and PD presents evidence that module-level analysis may capture shared biology that individual proteins cannot.

Throughout this study, we primarily defined two protein sets, those selected by LASSO shrinkage in the ProRS model and those significantly predicted by PRS, and we showed that the two are largely disjoint. However, the *L*_1_ penalty has no grouping effect. Among correlated predictors the LASSO retains a subset and shrinks the rest to zero, and which member survives may vary across resamples, so a protein that is both PRS-associated and predictive could be dropped in favor of a correlated partner, deflating the overlap.^39,40^ The elastic-net refit rules this out. Admitting correlated groups together decreased the ratio instead of the expected increase, so the separation is a property of the two sets rather than an artifact of how one was selected. For future studies, the two sets may be seen as complementary rather than mutually-replaceable, and provide maximal signal when they converge. CR1, for example, is shared by both sets in AD, and external Mendelian randomization work has supported its causal role.^41,42^

Finally, every estimate rests on very small polygenic signal once *APOE* is removed, where the scores explain under 1% of liability and the three remaining absorption intervals reach lower bounds of 0.12, 0.01 and 0.00. Larger discovery GWAS would resolve this, and phenotype definition compounds the problem. The FTD disase model is uninterpretable at the available GWAS size and was excluded from the inferential table rather than reported as null, and amyotrophic lateral sclerosis cases were scored as a single trait without stratification on *C9orf72* or *SOD1*, so a heterogeneous liability was treated as homogeneous. Genotype-stratified replication in a consortium cohort would separate these.

## Data Availability

The data supporting these findings are available through the UK Biobank (ukbiobank.ac.uk) and the All of Us Research Program (researchallofus.org) to authorized researchers upon approval of an access application.

https://github.com/czbw022/Absorption-and-Co-expression-Modules-Show-Where-PRS-ProRS-Diverge-in-Neurodegenerative-Diseases

## Acknowledgments

This work was supported by the National Institute of Health U01 AG068057.

## Supplementary Data

The supplementary files are available at https://github.com/czbw022/Absorption-and-Co-expression-Modules-Show-Where-PRS-ProRS-Diverge-in-Neurodegenerative-Diseases

